# Small Patient Datasets Reveal Genetic Drivers of Non-Small Cell Lung Cancer Subtypes Using a Novel Machine Learning Approach

**DOI:** 10.1101/2021.07.27.21261075

**Authors:** Cook Moses, Qorri Bessi, Baskar Amruth, Ziauddin Jalal, Pani Luca, Yenkanchi Shashibushan, Joseph Geraci

## Abstract

**Background:** There are many small datasets of significant value in the medical space that are being underutilized. Due to the heterogeneity of complex disorders found in oncology, systems capable of discovering patient subpopulations while elucidating etiologies is of great value as it can indicate leads for innovative drug discovery and development.

**Materials and Methods:** Here, we report on a machine intelligence-based study that utilized a combination of two small non-small cell lung cancer (NSCLC) datasets consisting of 58 samples of adenocarcinoma (ADC) and squamous cell carcinoma (SCC) and 45 samples (GSE18842). Utilizing a set of standard machine learning (ML) methods which are described in this paper, we were able to uncover subpopulations of ADC and SCC while simultaneously extracting which genes, in combination, were significantly involved in defining the subpopulations. We also utilized a proprietary interactive hypothesis-generating method designed to work with machine learning methods, which provided us with an alternative way of pinpointing the most important combination of variables. The discovered gene expression variables were used to train ML models. This allowed us to create methods using standard methods and to also validate our in-house methods for heterogeneous patient populations, as is often found in oncology.

**Results:** Using these methods, we were able to uncover genes implicated by other methods and accurately discover known subpopulations without being asked, such as different levels of aggressiveness within the SCC and ADC subtypes. Furthermore, *PIGX* was a novel gene implicated in this study that warrants further study due to its role in breast cancer proliferation.

**Conclusion:** Here we demonstrate the ability to learn from small datasets and reveal well-established properties of NSCLC. This demonstrates the utility for machine learning techniques to reveal potential genes of interest, even from small data sets, and thus the driving factors behind subpopulations of patients.

## 1. Introduction

The collection of transcriptomic data is expensive, resulting in datasets with a small number of sample sizes (in the hundreds) but thousands of variables. As a result, several techniques that are making significant strides in the imaging space, such as deep neural networks, are not suitable for these datasets, as a large number of samples are required. Furthermore, the heterogeneity of the patient population and the complexity of diseases found in oncology requires going beyond the labels. The development of techniques that can explain the driving variables behind patient subpopulations is tremendously valuable in identifying and developing novel therapeutic agents – this is particularly relevant for mapping out heterogeneous diseases such as lung cancer.

Lung cancer is the leading cause of cancer mortality worldwide, with non-small cell lung cancer (NSCLC) accounting for 85% of all lung cancers [1]. NSCLC can be divided into three histological subtypes with distinct phenotypes and prognoses: adenocarcinoma (ADC), squamous cell carcinoma (SCC) and large cell carcinoma (LCC) [2, 3]. The histological differences across these subtypes suggest that distinct molecular mechanisms underlie the observed phenotypic differences. Although the differential gene expressions across NSCLC subtypes have been of increasing interest, the therapeutic implications on how these pathways interact, is only more recently being investigated [4]. The remarkable degree of genetic variability within each histological subtype only highlights the importance of molecular biology and genotyping for NSCLC [5, 6].

Fortunately, machine learning (ML) advancements have served as promising tools for stratifying NSCLC, predicting transcriptional mutations based on histological slides or discriminating NSCLC subtype through genomic expression levels. The bulk of ML efforts have focused on image analysis for predicting the stage of NSCLC [7-10]. However, the growing body of evidence highlighting the molecular abnormalities that underlie the genomic subtypes of NSCLC can train ML algorithms to identify novel biomarkers for NSCLC, moving towards precision medicine [11-13]. For instance, previous reports have identified that ADC is associated with increased expression of genes related to protein transport and cell junctions, while SCC is associated with increased expression of genes related to cell division and DNA replication [14]. An analysis of gene expression profiles between ADC and SCC using machine learning has been previously reported, identifying several genes including *CSTA, TP63, SERPINB13, CLCA2, BICD2, PERP, FAT2, BNC1, ATP11B, FAM83B, KRT5, PARD6G*, and *PKP1* which were differentially expressed in ADC and SCC [15].

Here, using a combination of ML tools designed to learn from patient datasets to analyze gene expression data derived from ADC and SCC NSCLC patients, we were able to identify novel driving genes that distinguish these two broad subtypes. ML with statistical modelling tailored for small datasets has shown promise in showcasing disease heterogeneity[16]. Because large datasets are critical for contemporary machine learning methods such as CNNs, there is a need for alternative techniques when data banks are insufficient to train the model. In addition, significant features found within small datasets may become diluted by more obvious statistical features and hence over-represented in large datasets. As such, ML methods must be carefully used and complemented by statistical methods that allow for the discovery of non-linear ways in which groups of genes may interact to drive disease heterogeneity. The methodology presented here is designed for small datasets, which presents as a novel way of hypothesizing genetic subpopulations that may result in pathanogenesis. Our findings support genes previously reported to distinguish ADC and SCC subtypes. However, the novelty of this work lies in the machine’s ability to discover previously unknown subpopulations that are defined by several genes at a time. These findings shed light on the different mechanisms at play within these subtypes.

This article has been formatted according to the TRIPOD guidelines.

## 2. Materials and Methods

### Datasets

The dataset consisted of 40 samples of ADC and 18 samples of SCC (GSE10245) [17] and 9 samples of ADC and 36 samples of SCC (GSE18842) [18] to obtain a total of 103 samples. Only GSE10245 was used when analyzing gene expression levels for discriminating differences between sex as this data was omitted from GSE18842. Genetic expression levels denote relative RMA-calculated signal intensity [19]. Bar plot means represent the mean expression level and error bars represent the standard deviation of the pooled data from each probe ID.

We utilized publicly available data sets that upon inspection had excellent signal for separating out adenocarcinoma and squamous cell carcinoma. The data consists of gene expression and is very expensive to acquire. We decided to analyze these two data sets because we were interested in what we could accomplish with a smaller than ideal data set using machine learning. This paper is a report of our findings after using a set of techniques that are appropriate for small data in order to encourage others to explore smaller data sets as there may be hidden valuable information within them that could be extracted with the techniques we described.

### Machine Intelligence

In this study, we used a methodology to organize the resulting models from several well-known machine learning methods to explore NSCLC genetic heterogeneity within a small dataset. This organizational technique was used to extract insights from models that could then be compared with statistical methods suitable for small data. The only proprietary method used for these results are the techniques referred to as a feature selection tool [20, 21], in order to help us reduce the size of the data set to 16 dimensions. More specifically, we used these methods to create several new 16-dimensional data sets. We then used the following algorithm, based on standard methods, to create models and insights. For the work reported in this paper, we utilized the following process, after we performed our feature reduction:

1. First, a simple variable reduction was performed via standard univariate reduction methods and ensemble trees (Random Forest) through cross-validation [22, 23]. The only dependent variables used were ADC vs SCC. All univariate statistical methods incorporated Bonferroni corrections.
2. At this point we exercised two options: a) we used methods [20] to arrive at 16 variable data sets (in order to test this system), and b) we allowed step 1 above to be our sole variable selection method. For replication purposes, one may run step 1 alone.
3. Principal components were utilized as a linear unsupervised clustering method to reveal obvious subpopulation structures.
4. The loadings from the principal components were utilized to reduce the variables.
5. Using the t-SNE [24], HDBSCAN [25] and UMAP [26] algorithms, we were able to extract subpopulations.
6. We then collected the sample IDs from the clusters formed from these two clustering models, systematically compared each group with the others, and then applied statistical methods to determine differentially expressed gene candidates.
7. In order to determine the significance of a gene, a standard Student t-test was used when two subpopulations were compared, and if more than two subpopulations were compared, then an ANOVA was used. We then plotted the resulting clusters for the purpose of illustrating our findings. Again, these methods incorporated Bonferroni corrections.

Clustering was performed via principal components, t-SNE, HDBSCAN and UMAP and these were the basis of the maps found in this paper. Some proprietary algorithms were used to organize the resulting clustering models, in addition to the random forest models, such that we were able to explore the models interactively to derive a deeper understanding of the driving genes behind the sub-clusters [20]. The NetraAI system goes beyond these capabilities, but we did not utilize these proprietary methods to maintain academic standards. By allowing ourselves to use the proprietary organization methods provided by the NetraAI, we were able to identify subpopulations that we could compare with statistical methods suitable for a dataset with so few samples and avoid overfitting that often comes with utilizing machine learning methods with small datasets.

## 3. Results

### 3.1 Machine learning identifies differentially expressed genes from a small NSCLC dataset

Using the ADC and SCC tumor gene expression data, our approach was able to generate a map distinguishing SCC (blue) and ADC subjects (red) (Figure 1). The genes that were found to have driven this distinction were *DSC3, VSNL1, SLC6A10P, IRF6, DST, CLCA2, DSG3, LPCAT1* and *PIGX*. Previous studies have reported on differentially expressed genes in ADC and SCC. Here, we identified 17 genes that discriminate between SCC and ADC (Table 1). It is noteworthy that 16 of the 17 genes we identified have been previously reported to be differentially expressed in SCC and ADC, validating our methods. Interestingly, we found genes associated with gap junctions and tight junctions to be strong driving forces differentiating SCC and ADC. It is noteworthy that *PIGX* was the only gene identified that has not been previously associated with NSCLC. Although there have been reports that *PIGX* promotes cancer cell proliferation by suppressing *EHD2* and *ZIC1*, this warrants further investigation [27].

**Figure 1.**
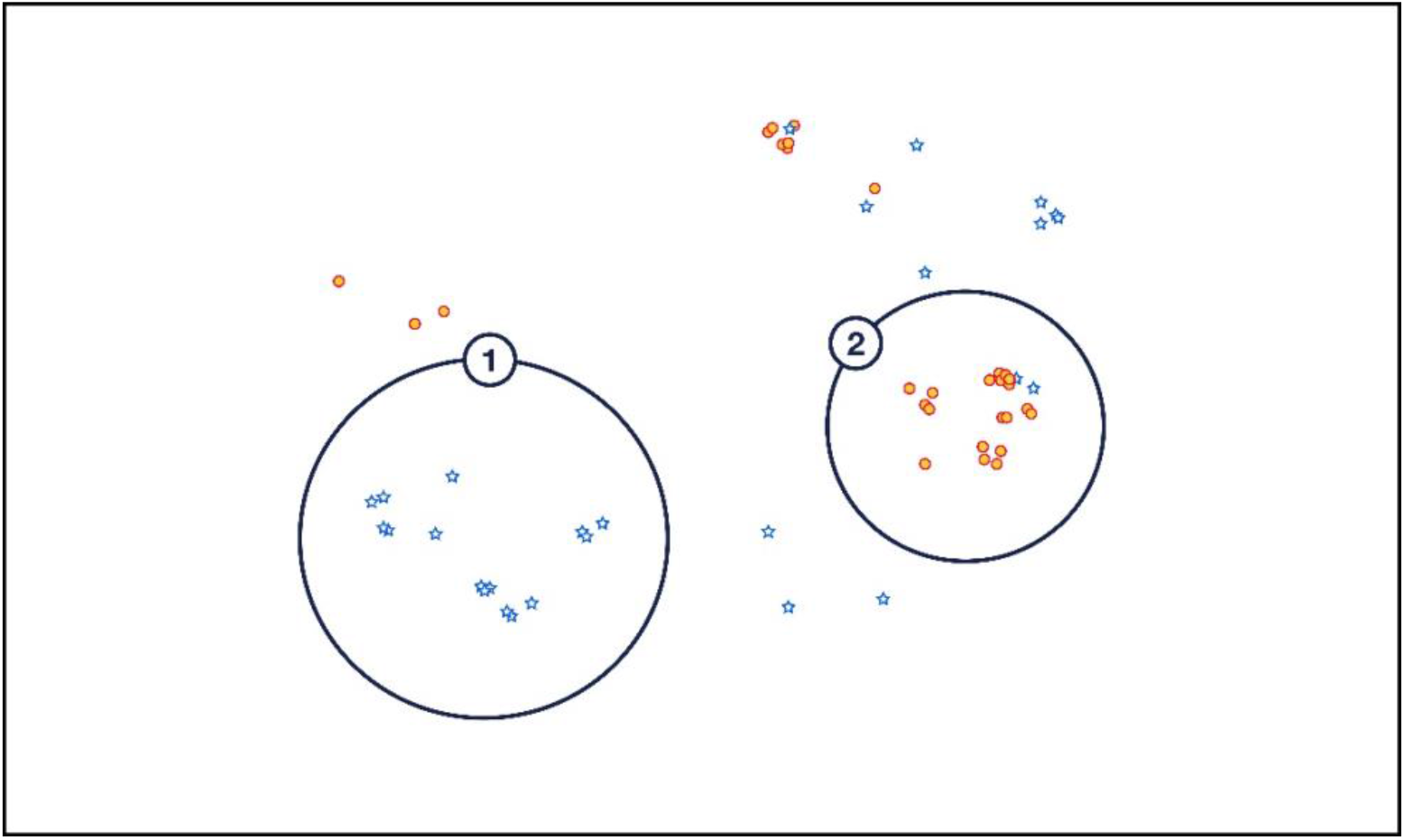
Classification of NSCLC patients stratified into SCC and ADC. SCC (blue) and ADC subjects (red) were delineated by HDBSCAN. The encircled groups were characterized by different expression levels of DSC3, VSNL1, SLC6A10P, IRF6, DST, CLCA2, DSG3, LPCAT1 and PIGX.

**Table 1.**
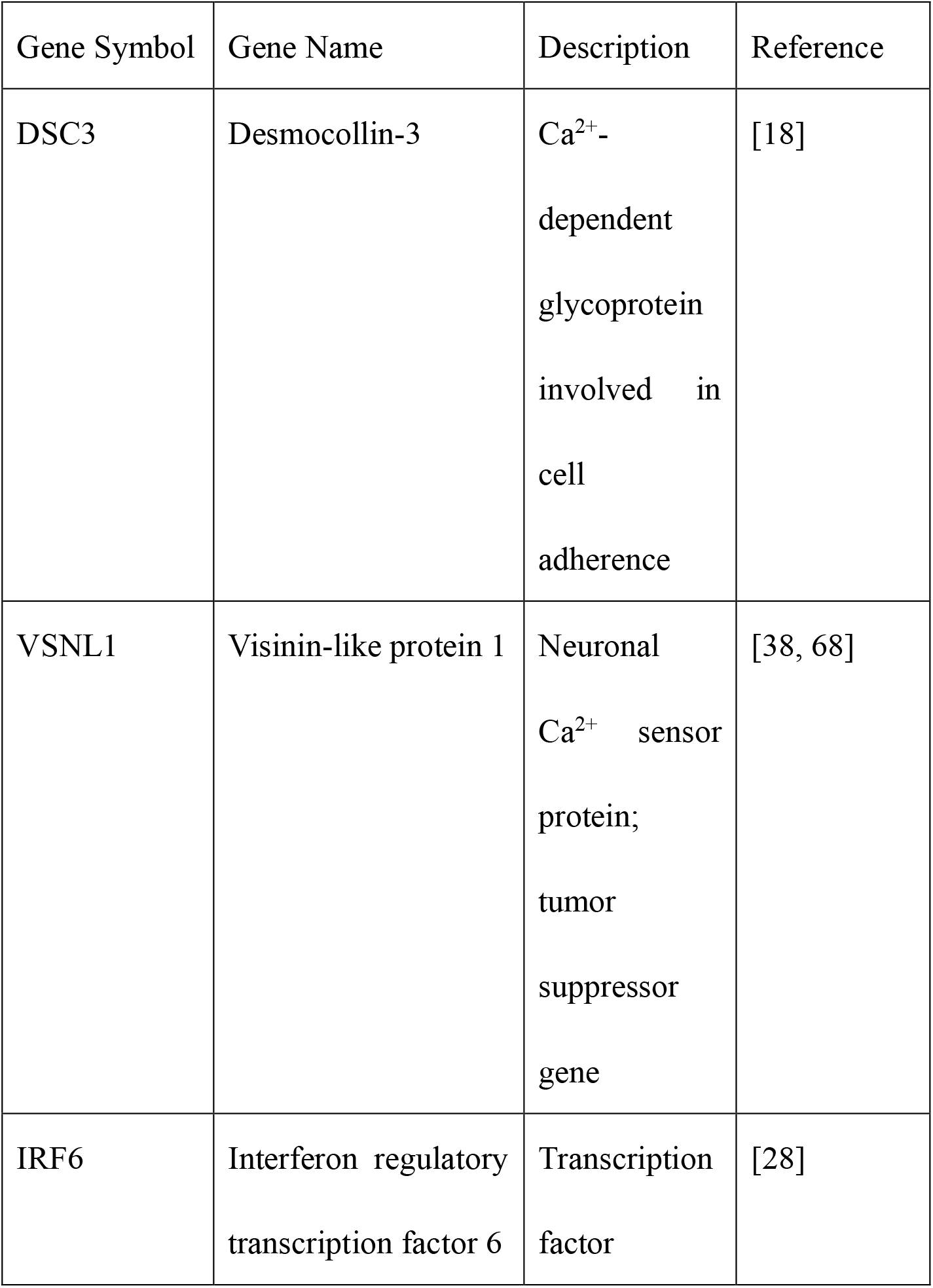
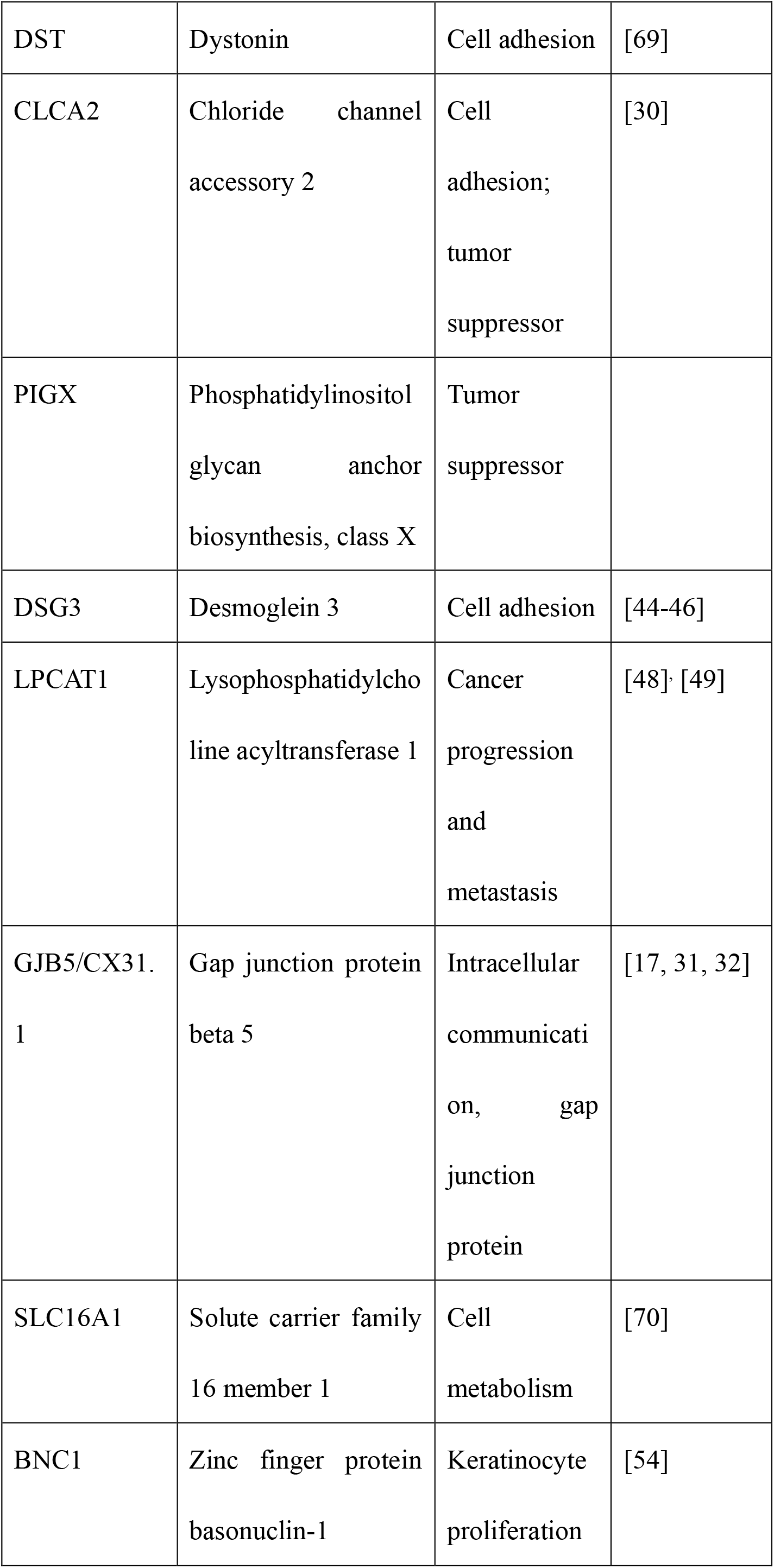
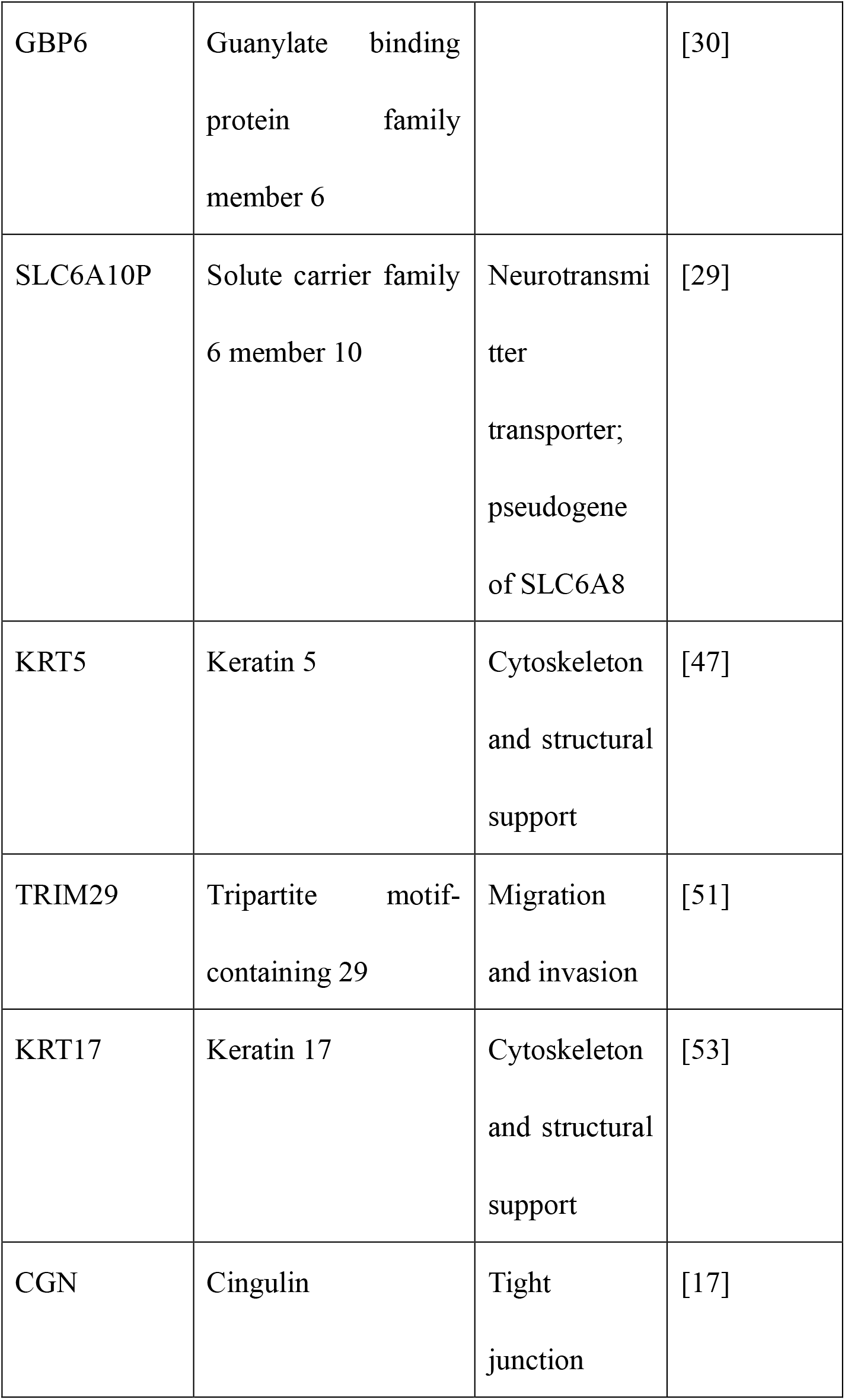
Genes identified as discriminating between squamous cell carcinoma and adenocarcinoma.

### 3.2 ADC and SCC are associated with distinct cellular adhesion molecules

Reports of SCC being characterized by the upregulation of desmosome and gap junction genes and ADC characterized by the upregulation of tight junction genes suggest that NSCLC subtypes are associated with a distinct set of adhesion molecules [17]. Here, we found that SCC was associated with cell adhesion marker *DSC3*, and ADC was associated with tight junction marker *CGN* (Figure 2). We identified two probes corresponding to *DSC3*, 206032_at and 206033_s_at. There was a statistically significant association of both *DSC3* probes with SCC (p < 0.0001) (Figure 2A). Interestingly, the elevated expression of *DSC3* was associated with males; however, this was not statistically significant (p = 0.062 for 206032_at and p = 0.077 for 206033_s_at). In contrast, the two probes corresponding to *CGN*, 223232_s_at and 223233_s_at were significantly associated with ADC (p < 0.0001) (Figure 2). The *CGN* probes were significantly associated with females (p = 0.014). The variability of adhesion molecule expression across sex warrants further investigation to elucidate the details of the correlation and advance towards gender related precision medicine.

**Figure 2.**
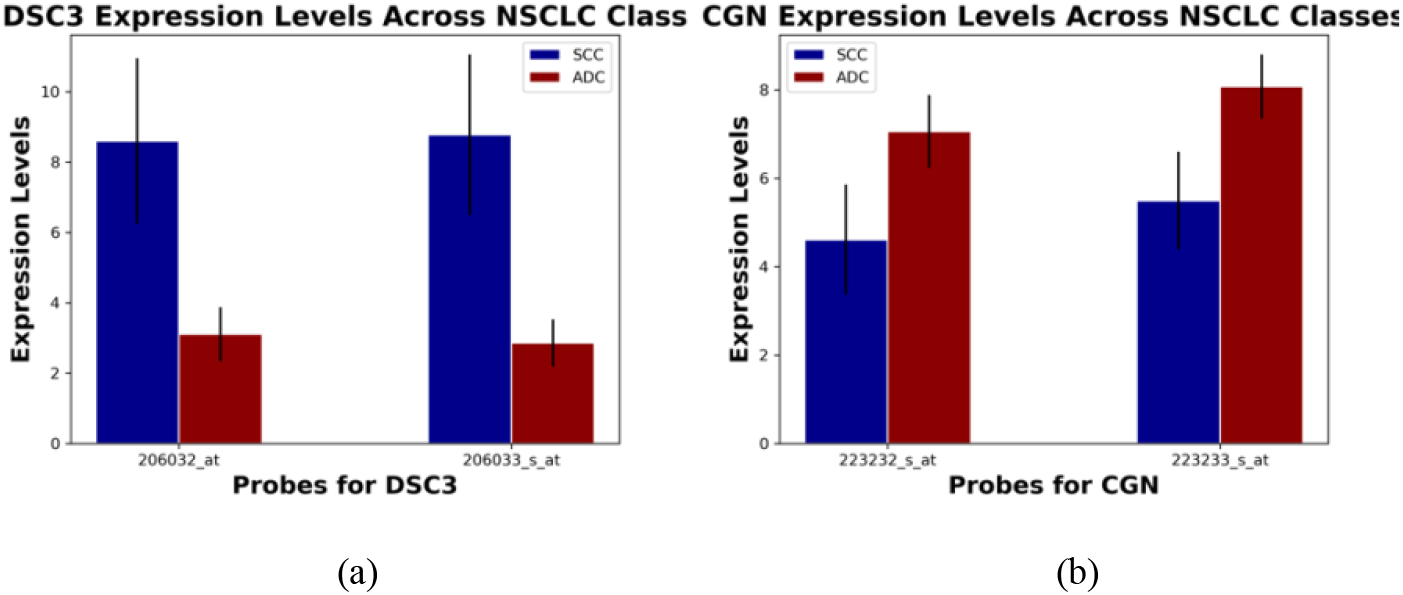
Differential expression of DSC3 and CGN in NSCLC. (A) Expression level of DSC3 probes (mean ± SD), 206032_at and 206033_s_at, in SCC and ADC. (B) Expression level of CGN probes, 223232_s_at and 223233_s_at, in SCC and ADC. Abbreviations: NSCLC, non-small cell lung cancer; SD, standard deviation; SCC, squamous cell carcinoma; ADC, adenocarcinoma.

### 3.3 SLC6A10P may be a key driver of a more aggressive ADC subtype

Elevated expression of *SLC6A10P* was significantly associated with two subgroups of ADC (p<0.0001) (Figure 3), in line with previous reports [28, 29]. Interestingly, increased expression of the pseudogene *SLC6A10P* in ADC has been associated with increased metastatic risk and reported to be a significant predictor of poor clinical outcome [29]. Our ML methodology was able to reveal subpopulations of ADC subjects that are uniquely classified by *SLC6A10P* (p = 1.3×10^−5^).. This demonstrates the potential power of machine intelligence to reveal aetiologias within complex diseases, even when a small number of samples are present. However, the methods must be used to reveal subpopulations that can then be compared using appropriate statistical methods suitable for comparing small groups.

**Figure 3.**
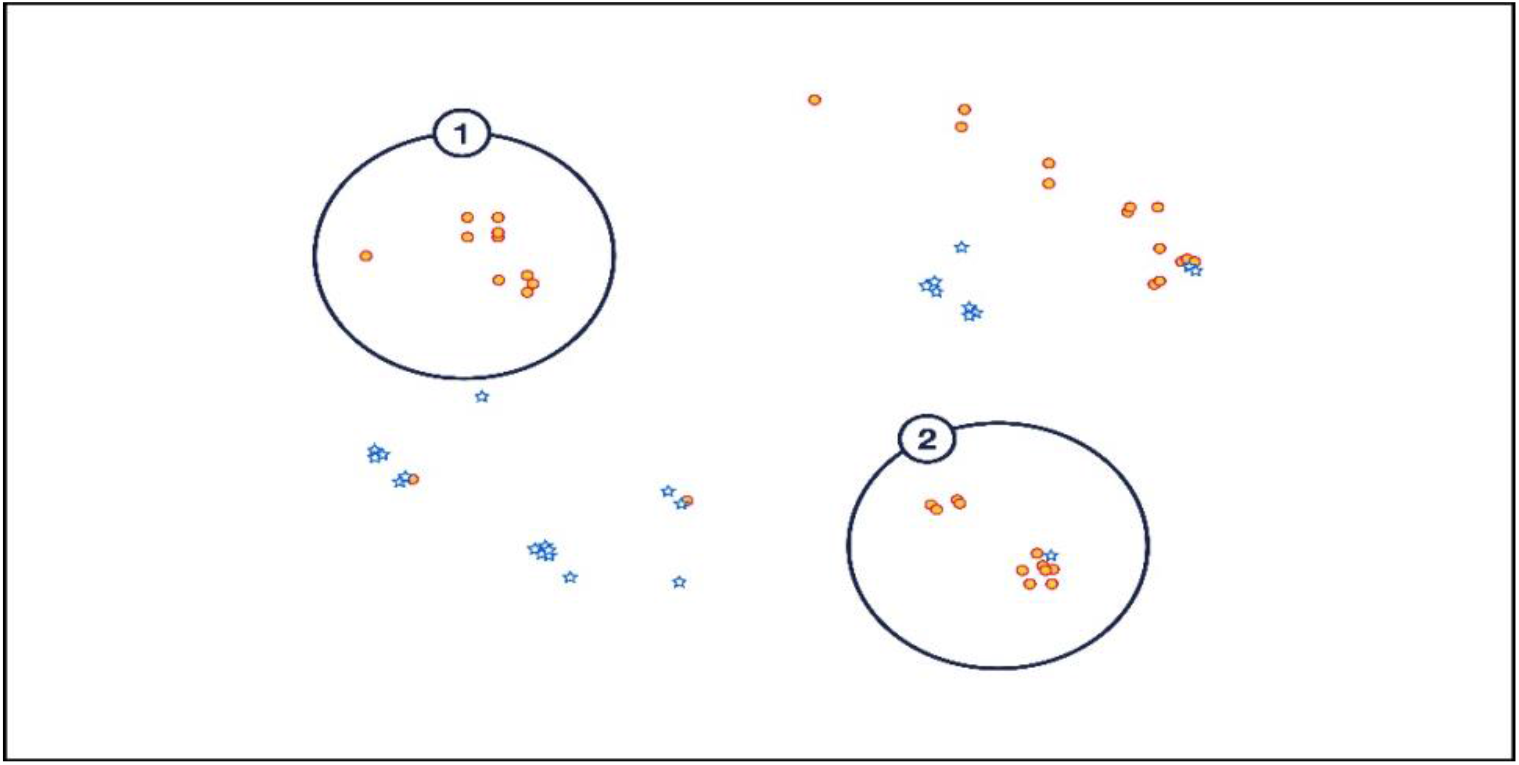
Unsupervised clustering of ADC subject subgroups. ADC (adenocarcinoma) subject (red) subgroups were delineated via the aforementioned clustering methods, namely HDBSCAN. These two ADC patient subgroups are driven by SLC6A10P.

### 3.4 IRF6 and CLCA2 drive unique subpopulations of SCC

Consistent with previous reports, we found two distinct subpopulations of SCC were found to be driven by *IRF6* and *CLCA2* (Figure 4) [28, 30]. *IRF6* and *CLCA2* expression levels were higher in SCC than ADC (p<0.0001) (Figure 4B and 4C). The significance value between the *CLCA2* and *IRF6* probes in the two encircled SCC groups were evaluated to be 4.4×10^−7^, 5.8×10^−3^, 9.3×10^−7^ and 0.046 for the 206164_at, 206165_s_at, 206166_s_at and 1552477_a_at probes, respectively.

**Figure 4.**
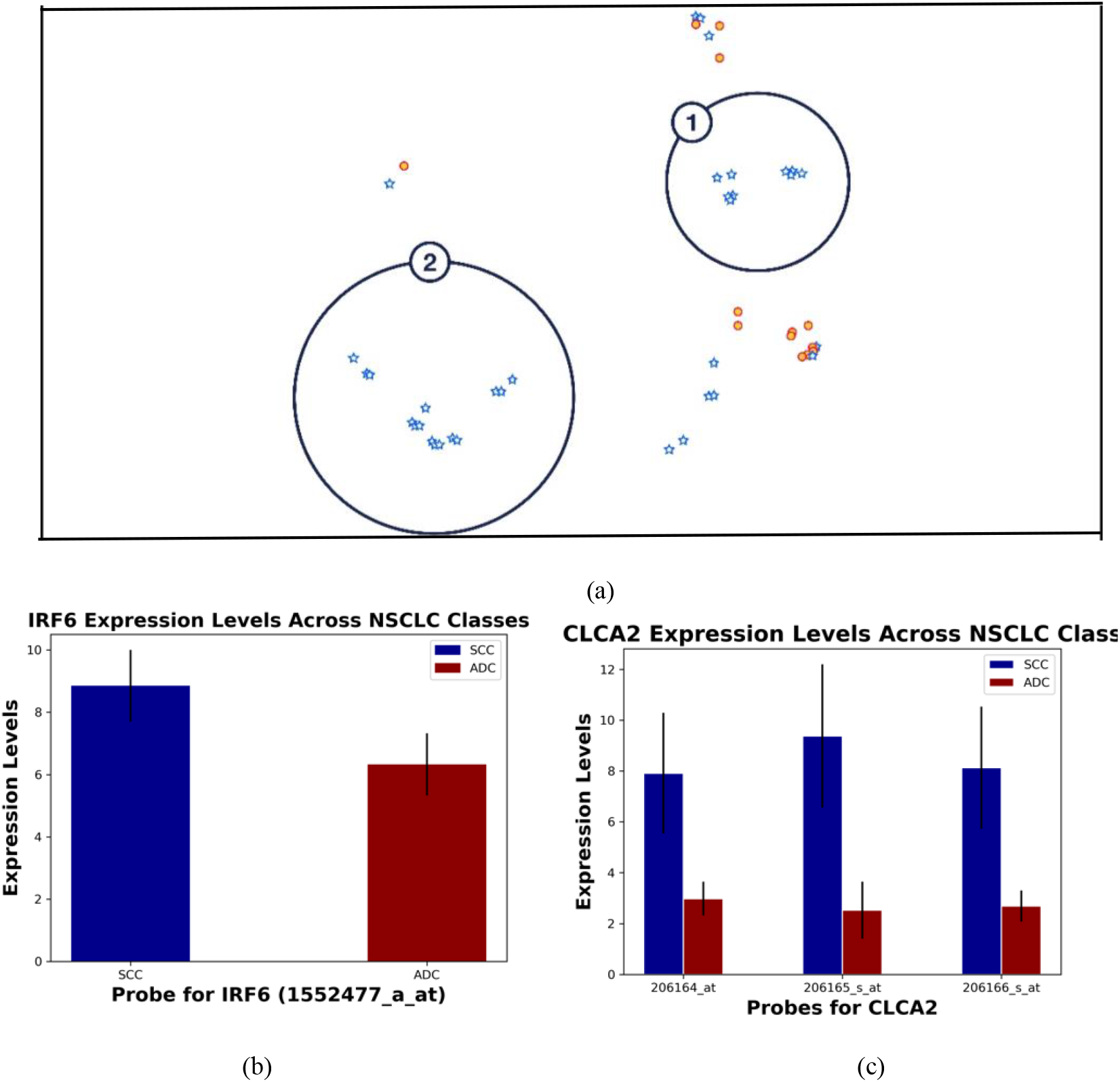
Differential expression of IRF6 and CLCA2 in NSCLC. (A) Our methodology hypothesized that the encircled SCC (squamous cell carcinoma) patient subgroups were driven by IRF6 and CLCA2. (B) Expression level of the IRF6 probe (mean ± SD), 1552477_a_at in SCC and ADC. (C) Expression level of CLCA2 probes 206164_at, 206165_s_at, and 206166_s_at in SCC and ADC (mean ± SD).

## 4. Discussion

This study highlights the genetic heterogeneity within NSCLC subtypes. Using a small dataset, we were able to identify a set of 17 genes that distinguish SCC and ADC (Table 1). Within these 17 genes, most have been previously reported to be associated either with NSCLC or a specific subtype of NSCLC, validating our ML approach. These findings were aligned with previous reports on SCC genes being associated with the organization and assembly of cell and gap junctions, glutathione conjugation and the redox stress response, ECM organization and collagen-related proteins, interferon and cytokine signaling, and HLA downregulation and ADC genes associated with ECM organization proteins and complement, interferon and cytokine signaling, and collagen-related genes and proteins for ECM organization [31]. Another study identified epidermis development, cell division, and epithelial cell differentiation as the most common categories characterizing SCC, and cell adhesion enrichment, biological adhesion, and coagulation for ADC [32]. However, some of the genes we identified have not been previously associated with NSCLC or a specific subtype and represent areas that warrant greater investigation for the advancement of precision medicine in NSCLC. Below, the genes of interest found using our methodology are highlighted in the context of previous findings in NSCLC.

The first of the previously reported NSCLC-associated genes we identified, *DSC3*, plays a role in epidermal morphology and keratinocyte proliferation [18]. There are several studies that report on *DSC3* distinguishing ADC and SCC, with a higher expression in SCC [33-36]. Notably, there has been a report on the association between *DSC3* and tumor suppressor activity in NSCLC mediated by inhibition of *EGFR* [37]. However, there remain contradictory associations with *DSC3* and prognosis, with elevated levels associated with increased metastatic risk in melanoma and better prognosis in lung and colon cancer [35]. This suggests that the same molecule may have differential effects in the tumor microenvironment (TME), which presents as an interesting field of research to understand how *DSC3* expression correlates with NSCLC subtypes depending on where they originate in the lung.

*VSNL1* codes for the calcium-sensor protein *VILIP1*. Lower *VSNL1* expression has been correlated with poor clinical outcomes in NSCLC patients [38]. *VILIP1* has been reported to be decreased or undetectable in aggressive and invasive SCC, while less aggressive SCC displayed *VILIP-1* expression [38]. There is evidence linking decreased *VILIP1* expression to increased cell motility and malignancy, suggesting that *VSNL1* downregulation promotes SCC tumor invasiveness [39].

Although a direct role of *IRF6* in lung cancer has not been identified, studies suggest that *IRF6* is a crucial regulator of the cell cycle, promoting progression to the G0 state and allowing for uncontrolled cell proliferation [18]. Decreased *IRF6* expression has been associated with poor prognosis of gastric cancer and increased invasiveness of breast cancer [40, 41].

Interestingly, *SLC6A10P* was the single gene that we found to drive two specific subtypes of ADC. *SLC6A10P* was previously found to be a marker for aggressive ADC [29], and recently, implicated within the Notch signaling pathway [42]. Our findings suggest that *SLC6A10P* warrants further investigation as a genetic biomarker in the context of the ADC patient subpopulation.

*DST* and *DSC3* have been increasingly reported to be highly expressed in both ADC and SCC. Overexpression of these desmosomal genes are associated with increased CD8^+^ T-cell infiltration in ADC [35].

*CLCA2* has been implicated as a negative regulator of cancer cell migration [43]. In the lung, *CLCA2* has been reported to be highly expressed in SCC, suggesting that it may serve as a diagnostic marker to differentiate SCC from ADC. Female patients with *CLCA2*-negative SCC exhibited significantly poorer prognoses [30].

*DSG3* has been reported to play a role in SCC and has been used as a sensitive and specific marker for SCC. It was also shown to be an effective discriminator between SCC and ADC [44, 45]. Higher *DSG3* expression correlated with lower survival in SCC [46]. *DSG3* and *KRT5* have been reported to be downregulated in AC [47].

*LPCAT1* has recently been shown to be overexpressed in lung SCC and associated with decreased OS [48]. In lung ADC, gene overexpression was associated with higher probabilities of ADC metastasis and poor clinical outcomes [49].

There is evidence that supports the role of cell adhesion proteins in both ADC and SCC. However, *GJB5* has been implicated in SCC mechanisms and is associated with gap junctions [31]. It is not surprising that there is a higher expression of *GJB5* in SCC as it is primarily associated with gap junctions (Figure 2) [17, 32]. *GJB5* (gap junction protein beta 5 or protein-coding gene: *Cx31*.*1*) is involved in intercellular communication related to epidermal differentiation and environmental sensing.

*Cx31*.*1* was found to be downregulated in NSCLC with expression levels inversely related to metastatic potential, suggesting it inhibits malignant properties of NSCLC cell lines. *Cx31*.*1* is colocalized with LC3-II (autophagy marker light chain 3) and acts as a tumor suppressor as it plays a role in the regulation of cell proliferation, cell differentiation, tissue development and apoptosis [32].

*TRIM29* has been shown to be upregulated in NSCLC, and may be a marker for tumour aggressiveness [50]. It has been further associated with poorer histological grade and clinical outcomes in SCC [51]. It has been suggested that this may be due to the inhibition of p53 via *TRIM29* [52].

*KRT17* overexpression has been associated with both subtypes of NSCLC, but was significantly correlated to more advanced tumour grade, lymph node metastatic potential, and overall survival in ADC [53].

*BCN1* has been reported to be hypermethylated in NSCLC tissue [54]. Furthermore, decreased expression of *BNC1* has been observed in other carcinomas [55]. Aberrant *BNC1* and *BNC2* expression contributes to tumor progression [56].

Reports of upregulation of desmosomes and gap junctions in SCC and tight junctions in ADC suggests that SCC and ADC are characterized by a distinct set of adhesion molecules [17]. Here, we found that ADC was identified by *CGN* and SCC by *DSC3* (Figure 2). *CGN* (cingulin) is involved in the organization of tight junctions and is downregulated in SCC [17]. In contrast, ADC has been reported to be characterized by tight junctions, while SCC is characterized by gap junctions.

In addition to the 17 identified genes differentially expressed in ADC and SCC, *PTGFRN* (prostaglandin F2 receptor negative regulator; CD315) was also found to be associated with ADC. *PTGFRN* has been reported to be associated with worse survival in glioblastoma, while inhibition has been associated with decreased proliferation and tumor growth [57, 58]. *PTGFRN* inhibits the binding of prostaglandin F2α to its receptor. Notably, there are reports that *PTGFRN* is associated with small cell lung cancer; however, the role remains unknown [59, 60].

*IRF6* and *CLCA2* have previously been implicated in lung SCC [28, 30]. *CLCA2* in particular was highlighted to differentiate ADC and SCC. Furthermore, SCC expression was correlated with tumour grade upon histological characterization. In particular, *CLCA2* negative samples were associated with poorly differentiated tumours [30].

Males have been reported to have a significantly poorer NSCLC prognosis compared to females, shifting efforts towards sex-based approaches to diagnosis, prognosis, and therapeutic interventions [61, 62]. Additionally, estrogens have been associated with increased risk in ADC in women despite equal expression of estrogen receptors α and β, however, the role remains unclear [63]. While there are several reports on the sex-based differences in cancer mechanisms, including differences in metabolism, immunity, and angiogenesis, differences in *CGN* and *DSC3* expression have not been previously reported to the best of our knowledge [64]. Gap junction proteins, also known as connexins, serve as channels that connect the interior of adjacent cells, facilitating intracellular homeostasis and coordination of activities via second messengers [65]. Desmosomes primarily provide mechanical strength via a structural network. In contrast, tight junctions form a barrier around the cell, regulating permeability of the paracellular space [66, 67]. These molecules play critical roles in epithelial-to-mesenchymal transition, a process involved in cancer metastasis. Though no sex-based differences have been reported, this presents as a unique field of research, as there may be different druggable targets for males and females.

Finally, the phosphatidylinositol glycan anchor biosynthesis class gene, *PIGX*, was found to be a driver of ADC and SCC differentiation in several instances (Figure 1). Little is known about the role of *PIGX* in NSCLC. However, it has been noted that *PIGX* has a proliferative role when expressed in breast cancer cells [27]. In addition, authors found higher *PIGX* expression was associated with shorter recurrence-free survival. This suggests that this gene plays a role in NSCLC that warrants further study.

## 5. Conclusions

The approach utilized here to derive the insights relied on the ability for certain machine learning methods to create hypotheses about subpopulations of patients, and then to statistically test the driving variables of these subgroups of patients. In this way, we utilize machine learning to derive potential insights and then utilize statistical methods that are suitable for small data to evaluate differential expression. In order to create robust predictive models with machine intelligence, one requires large data sets, but here we utilized the ability for some of these methods to create hypotheses instead, and then use methods appropriate for small data to test these hypotheses. This bidirectional attack allowed us to derive insights from these small datasets that have been previously validated and to finally derive a new potential role for the gene *PIGX* in NSCLC.

### Limitations

This study highlights a methodology that targets genes of interest in a small, heterogenous subpopulation of NSCLC. Although small populations are more prone to local drivers of genetic heterogeneity, they will not encompass all genes that may drive other subtypes of NSCLC in patients. Therefore, a limitation of this methodology resides in it’s inability to forecast more obvious patterns found in larger datasets.

## Data Availability

All data was procured from a publicly available database from the Gene Expression Omnibus. The dataset consisted of 58 samples of ADC and SCC (GSE10245) and 45 samples of human lung cancer and controls (GSE18842) to obtain a total of 103 samples.

https://www.ncbi.nlm.nih.gov/geo/query/acc.cgi?acc=GSE10245

## Abbreviations

ADC: adenocarcinoma
AUC: area under the curve
CNN: convolutional neural network
CT: computed tomography
EMT: epithelial-to-mesenchymal transition
LCC: large cell carcinoma
ML: machine learning
NSCLC: non-small cell lung cancer
PET: positron emission tomography
ROC: receiver operator curve
SCC: squamous cell carcinoma
SVM: support vector machine
TME: tumor microenvironment

## Footnote

The TRIPOD reporting statement has been completed.

The authors are accountable for all aspects of the work in ensuring that questions related to the accuracy or integrity of any part of the work are appropriately investigated and resolved.

According to the TRIPOD Checklist: Prediction Model Development, the following Items can be found:

1. Page 1

2. Page 2

3a. Introduction. Page 4, paragraph 1

3b. Introduction. Page 4, paragraph 2

4a. Datasets: Page 5, paragraph 1

4b. Not applicable

5. Datasets: Page 5, paragraph 1

6. Machine Intelligence. Page 6.

7. Machine Intelligence. Page 6, 7.

8. Datasets. Page 6, paragraph 2

9. Complete case analysis

10 a. Machine Intelligence. Page 6, 7.

10 b. Machine Intelligence. Page 6, 7.

10 c. Machine Intelligence. Pages 6-8

11. Not applicable

13 a, b. Datasets: Page 5, paragraph 1

14 a. Datasets: Page 5, paragraph 1

14 b. Not applicable

15 a, b. Machine Intelligence. Pages 6-8

16. Results, Pages 8-16

18. Conclusion. Page 23

19b. Discussion. Page 16-21

20. Discussion. Page 22

21. Not applicable

22. Page 27

## Conflicts of interest

J.G. is a major shareholder of NetraMark Corp, where NetraMark is a technology company providing clinical trial support to pharmaceutical companies.

L.P. has previously acted as a scientific consultant for AbbVie USA; Acadia USA; BCG Switzerland; Boehringer Ingelheim International GmbH; Compass Pathways; EDRA-Publishing, Italy; Ferrer Spain; Gedeon-Richter, Hungary; Inpeco SA, Switzerland; Johnson & Johnson USA; NeuroCog Trials USA; Novartis-Gene Therapies, Switzerland; Otsuka USA; Pfizer Global USA; PharmaMar Spain; Relmada Therapeutics USA; Takeda, USA; VeraSci, USA; Vifor Switzerland.

## Ethical approval

Not applicable

## Consent to participate

Not applicable

## Availability of data and materials

Data was obtained from publicly available datasets GSE10245 https://www.ncbi.nlm.nih.gov/geo/query/acc.cgi?acc=GSE10245 and GSE18842 https://www.ncbi.nlm.nih.gov/geo/query/acc.cgi?acc=GSE18842

## Funding

Part of this research was funded by NetraMark Corp in the form of salary for Dr. Joseph Geraci, and computational resources.

## Notes

### Author Declarations

This data is freely available through the Gene Expression Omnibus site and is anonymized. Thus for this project there was only an internal review conducted by Dr. Joseph Geraci and Jalal Ziauddin who are both employees at NetraMark corp.

### Summary of Updates

Figures have been revised for clarity. Introduction has been revised for brevity.

## References

[1] C. A. Ridge, A. M. McErlean, and M. S. Ginsberg, “Epidemiology of lung cancer,” in Seminars in interventional radiology, 2013, vol. 30, no. 2: Thieme Medical Publishers, p. 93.

[2] A. Thomas, S. V. Liu, D. S. Subramaniam, and G. Giaccone, “Refining the treatment of NSCLC according to histological and molecular subtypes,” Nature reviews Clinical oncology, vol. 12, no. 9, p. 511, 2015.

[3] M. S. Lawrence et al., “Discovery and saturation analysis of cancer genes across 21 tumour types,” Nature, vol. 505, no. 7484, pp. 495–501, 2014.

[4] L. A. Pikor, V. R. Ramnarine, S. Lam, and W. L. Lam, “Genetic alterations defining NSCLC subtypes and their therapeutic implications,” Lung cancer, vol. 82, no. 2, pp. 179–189, 2013.

[5] C. Manegold, “Treatment algorithm in 2014 for advanced non-small cell lung cancer: therapy selection by tumour histology and molecular biology,” Advances in medical sciences, vol. 59, no. 2, pp. 308–313, 2014.

[6] S. Carnio, S. Novello, P. Bironzo, and G. V. Scagliotti, “Moving from histological subtyping to molecular characterization: new treatment opportunities in advanced non-small-cell lung cancer,” Expert review of anticancer therapy, vol. 14, no. 12, pp. 1495–1513, 2014.

[7] L. Yu et al., “Prediction of pathologic stage in non-small cell lung cancer using machine learning algorithm based on CT image feature analysis,” BMC cancer, vol. 19, no. 1, pp. 1–12, 2019.

[8] N. Tau, A. Stundzia, K. Yasufuku, D. Hussey, and U. Metser, “Convolutional neural networks in predicting nodal and distant metastatic potential of newly diagnosed non–small cell lung cancer on FDG PET images,” American Journal of Roentgenology, vol. 215, no. 1, pp. 192–197, 2020.

[9] M. Kriegsmann et al., “Deep Learning for the Classification of Small-Cell and Non-Small-Cell Lung Cancer,” Cancers, vol. 12, no. 6, p. 1604, 2020.

[10] W. Mu et al., “Non-invasive decision support for NSCLC treatment using PET/CT radiomics,” Nature communications, vol. 11, no. 1, pp. 1–11, 2020.

[11] M. Rabbani, J. Kanevsky, K. Kafi, F. Chandelier, and F. J. Giles, “Role of artificial intelligence in the care of patients with nonsmall cell lung cancer,” European journal of clinical investigation, vol. 48, no. 4, p. e12901, 2018.

[12] M. S. Lawrence et al., “Mutational heterogeneity in cancer and the search for new cancer-associated genes,” Nature, vol. 499, no. 7457, pp. 214–218, 2013.

[13] M. D. Podolsky, A. A. Barchuk, V. I. Kuznetcov, N. F. Gusarova, V. S. Gaidukov, and S. A. Tarakanov, “Evaluation of machine learning algorithm utilization for lung cancer classification based on gene expression levels,” Asian Pacific Journal of Cancer Prevention, vol. 17, no. 2, pp. 835–838, 2016.

[14] J. Li, D. Li, X. Wei, and Y. Su, “In silico comparative genomic analysis of two non-small cell lung cancer subtypes and their potentials for cancer classification,” Cancer Genomics-Proteomics, vol. 11, no. 6, pp. 303–310, 2014.

[15] F. Yuan, L. Lu, and Q. Zou, “Analysis of gene expression profiles of lung cancer subtypes with machine learning algorithms,” Biochimica et Biophysica Acta Molecular Basis of Disease, vol. 1866, no. 8, p. 165822, 2020.

[16] G. A. Robinson et al., “Disease-associated and patient-specific immune cell signatures in juvenile-onset systemic lupus erythematosus: patient stratification using a machine-learning approach,” (in eng), Lancet Rheumatol, vol. 2, no. 8, pp. e485–e496, Aug 2020, doi: 10.1016/S2665-9913(20)30168-5.

[17] R. Kuner et al., “Global gene expression analysis reveals specific patterns of cell junctions in non-small cell lung cancer subtypes,” Lung cancer, vol. 63, no. 1, pp. 32–38, 2009.

[18] A. Sanchez - Palencia et al., “Gene expression profiling reveals novel biomarkers in nonsmall cell lung cancer,” International journal of cancer, vol. 129, no. 2, pp. 355–364, 2011.

[19] Z. Wu, R. A. Irizarry, R. Gentleman, F. Martinez-Murillo, and F. Spencer, “A Model-Based Background Adjustment for Oligonucleotide Expression Arrays,” Journal of the American Statistical Association, vol. 99, no. 468, pp. 909–917, 2004/12/01 2004, doi: 10.1198/016214504000000683.

[20] B. Qorri, M. Tsay, A. Agrawal, R. Au, and J. Geraci, “Using machine intelligence to uncover Alzheimer’s disease progression heterogeneity,” (in en), Exploration of Medicine, vol. 1, no. 6, pp. 377–395, 2020/12/31/ 2020, doi: 10.37349/emed.2020.00026.

[21] M. Tsay, J. Geraci, and A. Agrawal, “Next-Gen AI for Disease Definition, Patient Stratification, and Placebo Effect,” OSF Preprints, 2020/04/06/T02:51:09.502Z 2020. Accessed: 2021/04/20/00:51:57. [Online]. Available: https://osf.io/pc7ak/

[22] C. Lai, M. J. Reinders, L. J. van’t Veer, and L. F. Wessels, “A comparison of univariate and multivariate gene selection techniques for classification of cancer datasets,” (in eng), BMC Bioinformatics, vol. 7, p. 235, May 2006, doi: 10.1186/1471-2105-7-235.

[23] X. Chen and H. Ishwaran, “Random forests for genomic data analysis,” (in eng), Genomics, vol. 99, no. 6, pp. 323–9, Jun 2012, doi: 10.1016/j.ygeno.2012.04.003.

[24] L. van der Maaten and G. Hinton, “Visualizing Data using t-SNE,” Journal of Machine Learning Research, vol. 9, pp. 2579--2605, 2008.

[25] M. H. Piekenbrock, Michael, “HDBSCAN with the dbscan package “, ed. https://cran.r-project.org/web/packages/dbscan/vignettes/hdbscan.html.

[26] L. McInnes, J. Healy, and J. Melville, “UMAP: Uniform Manifold Approximation and Projection for Dimension Reduction,” ed, 2018.

[27] M. Nakakido et al., “Phosphatidylinositol glycan anchor biosynthesis, class X containing complex promotes cancer cell proliferation through suppression of EHD2 and ZIC1, putative tumor suppressors,” International journal of oncology, vol. 49, no. 3, pp. 868–876, 2016.

[28] Y. Liu et al., “Interferon regulatory factor 6 correlates with the progression of non-small cell lung cancer and can be regulated by miR-320,” Journal of Pharmacy and Pharmacology, 2021, doi: 10.1093/jpp/rgab009.

[29] K. Yuan, Z.-J. Gao, W.-D. Yuan, J.-Q. Yuan, and Y. Wang, “High expression of SLC6A10P contributes to poor prognosis in lung adenocarcinoma,” International journal of clinical & experimental pathology, vol. 11, no. 2, p. 720, 2018.

[30] K. Shinmura et al., “CLCA2 as a Novel Immunohistochemical Marker for Differential Diagnosis of Squamous Cell Carcinoma from Adenocarcinoma of the Lung,” (in en), Disease Markers, Research Article 2014/12/07/ 2014.

[31] M. Lucchetta, I. da Piedade, M. Mounir, M. Vabistsevits, T. Terkelsen, and E. Papaleo, “Distinct signatures of lung cancer types: aberrant mucin O-glycosylation and compromised immune response,” BMC Cancer, vol. 19, no. 1, p. 824, 2019/08/20 2019, doi: 10.1186/s12885-019-5965-x.

[32] T. Wang, L. Zhang, P. Tian, and S. Tian, “Identification of differentially-expressed genes between early-stage adenocarcinoma and squamous cell carcinoma lung cancer using meta-analysis methods,” Oncology Letters, vol. 13, no. 5, pp. 3314–3322, 2017.

[33] A. Warth et al., “Large-scale comparative analyses of immunomarkers for diagnostic subtyping of non - small - cell lung cancer biopsies,” Histopathology, vol. 61, no. 6, pp. 1017–1025, 2012.

[34] K. Tsuta et al., “Utility of 10 immunohistochemical markers including novel markers (desmocollin-3, glypican 3, S100A2, S100A7, and Sox-2) for differential diagnosis of squamous cell carcinoma from adenocarcinoma of the Lung,” Journal of Thoracic Oncology, vol. 6, no. 7, pp. 1190–1199, 2011.

[35] Y. K. Chae et al., “Overexpression of adhesion molecules and barrier molecules is associated with differential infiltration of immune cells in non-small cell lung cancer,” Scientific reports, vol. 8, no. 1, pp. 1–10, 2018.

[36] B. Angulo et al., “Expression signatures in lung cancer reveal a profile for EGFR-mutant tumours and identify selective PIK3CA overexpression by gene amplification,” The Journal of pathology, vol. 214, no. 3, pp. 347–356, 2008.

[37] T. Cui et al., “The p53 target gene desmocollin 3 acts as a novel tumor suppressor through inhibiting EGFR/ERK pathway in human lung cancer,” Carcinogenesis, vol. 33, no. 12, pp. 2326–2333, 2012.

[38] J. Fu et al., “VILIP-1 downregulation in non-small cell lung carcinomas: mechanisms and prediction of survival,” PLoS One, vol. 3, no. 2, p. e1698, 2008.

[39] A. M. G. Guerrico, Z. M. Jaffer, R. E. Page, K.-H. Braunewell, J. Chernoff, and A. J. Klein-Szanto, “Visinin-like protein-1 is a potent inhibitor of cell adhesion and migration in squamous carcinoma cells,” Oncogene, vol. 24, no. 14, pp. 2307–2316, 2005.

[40] D. Li et al., “IRF6 is directly regulated by ZEB1 and ELF3, and predicts a favorable prognosis in gastric cancer,” Frontiers in oncology, vol. 9, p. 220, 2019.

[41] C. M. Bailey et al., “Mammary serine protease inhibitor (Maspin) binds directly to interferon regulatory factor 6: identification of a novel serpin partnership,” vol. 280, no. 40, pp. 34210–34217, 2005.

[42] Y. Feng, X. Guo, and H. Tang, “SLC6A8 is involved in the progression of non-small cell lung cancer through the Notch signaling pathway,” (in eng), Ann Transl Med, vol. 9, no. 3, p. 264, Feb 2021, doi: 10.21037/atm-20-5984.

[43] Y. Sasaki et al., “CLCA2, a target of the p53 family, negatively regulates cancer cell migration and invasion,” Cancer Biology & Therapy, vol. 13, no. 14, pp. 1512–1521, 2012/12/06/ 2012, doi: 10.4161/cbt.22280.

[44] J. Fukuoka et al., “Desmoglein 3 as a prognostic factor in lung cancer,” (in eng), Hum Pathol, vol. 38, no. 2, pp. 276–83, Feb 2007, doi: 10.1016/j.humpath.2006.08.006.

[45] C. D. Savci-Heijink et al., “The role of desmoglein-3 in the diagnosis of squamous cell carcinoma of the lung,” (in eng), Am J Pathol, vol. 174, no. 5, pp. 1629–37, May 2009, doi: 10.2353/ajpath.2009.080778.

[46] Y. Dong et al., “Desmoglein 3 and Keratin 14 for Distinguishing Between Lung Adenocarcinoma and Lung Squamous Cell Carcinoma,” (in eng), Onco Targets Ther, vol. 13, pp. 11111–11124, 2020, doi: 10.2147/OTT.S270398.

[47] J. Xiao et al., “Eight potential biomarkers for distinguishing between lung adenocarcinoma and squamous cell carcinoma,” (in eng), Oncotarget, vol. 8, no. 42, pp. 71759–71771, 2017, doi: 10.18632/oncotarget.17606.

[48] F. Liu et al., “A miR-205-LPCAT1 axis contributes to proliferation and progression in multiple cancers,” (in eng), Biochem Biophys Res Commun, vol. 527, no. 2, pp. 474–480, 06 2020, doi: 10.1016/j.bbrc.2020.04.071.

[49] C. Wei et al., “LPCAT1 promotes brain metastasis of lung adenocarcinoma by up-regulating PI3K/AKT/MYC pathway,” (in eng), J Exp Clin Cancer Res, vol. 38, no. 1, p. 95, Feb 2019, doi: 10.1186/s13046-019-1092-4.

[50] X. Song, C. Fu, X. Yang, D. Sun, X. Zhang, and J. Zhang, “Tripartite motif-containing 29 as a novel biomarker in non-small cell lung cancer,” (in eng), Oncol Lett, vol. 10, no. 4, pp. 2283–2288, Oct 2015, doi: 10.3892/ol.2015.3623.

[51] Z. Y. Zhou, G. Y. Yang, J. Zhou, and M. H. Yu, “Significance of TRIM29 and β-catenin expression in non-small-cell lung cancer,” (in eng), J Chin Med Assoc, vol. 75, no. 6, pp. 269–74, Jun 2012, doi: 10.1016/j.jcma.2012.04.015.

[52] Z. Yuan et al., “The ATDC (TRIM29) protein binds p53 and antagonizes p53-mediated functions,” (in eng), Mol Cell Biol, vol. 30, no. 12, pp. 3004–15, Jun 2010, doi: 10.1128/MCB.01023-09.

[53] Z. Wang et al., “Overexpression of KRT17 promotes proliferation and invasion of non-small cell lung cancer and indicates poor prognosis,” (in eng), Cancer Manag Res, vol. 11, pp. 7485–7497, 2019, doi: 10.2147/CMAR.S218926.

[54] D. S. Shames et al., “A genome-wide screen for promoter methylation in lung cancer identifies novel methylation markers for multiple malignancies,” (in eng), PLoS Med, vol. 3, no. 12, p. e486. Dec 2006, doi: 10.1371/journal.pmed.0030486.

[55] Y. Wu, X. Zhang, Y. Liu, F. Lu, and X. Chen, “Decreased Expression of BNC1 and BNC2 Is Associated with Genetic or Epigenetic Regulation in Hepatocellular Carcinoma,” (in eng), Int J Mol Sci, vol. 17, no. 2, Jan 2016, doi: 10.3390/ijms17020153.

[56] Y. Wu, X. Zhang, Y. Liu, F. Lu, and X. Chen, “Decreased expression of BNC1 and BNC2 is associated with genetic or epigenetic regulation in hepatocellular carcinoma,” International journal of molecular sciences, vol. 17, no. 2, p. 153, 2016.

[57] J. Marquez, J. Dong, C. Dong, C. Tian, and G. Serrero, “Identification of Prostaglandin F2 Receptor Negative Regulator (PTGFRN) as an internalizable target in cancer cells for antibody-drug conjugate development,” Plos one, vol. 16, no. 1, p. e0246197, 2021.

[58] B. Aguila et al., “The Ig superfamily protein PTGFRN coordinates survival signaling in glioblastoma multiforme,” Cancer letters, vol. 462, pp. 33–42, 2019.

[59] J. George et al., “Comprehensive genomic profiles of small cell lung cancer,” Nature, vol. 524, no. 7563, pp. 47–53, 2015.

[60] J. Krushkal et al., “Epigenome-wide DNA methylation analysis of small cell lung cancer cell lines suggests potential chemotherapy targets,” vol. 12, no. 1, pp. 1–28, 2020.

[61] Z. Wainer et al., “Sex-dependent staging in non–small-cell lung cancer; analysis of the effect of sex differences in the eighth edition of the Tumor, Node, Metastases Staging System,” vol. 19, no. 6, pp. e933–e944, 2018.

[62] C. Radkiewicz, P. W. Dickman, A. L. V. Johansson, G. Wagenius, G. Edgren, and M. Lambe, “Sex and survival in non-small cell lung cancer: A nationwide cohort study,” PLoS ONE, vol. 14, no. 6, p. e0219206, 2019.

[63] M. M. Ivanova, W. Mazhawidza, S. M. Dougherty, and C. M. Klinge, “Sex differences in estrogen receptor subcellular location and activity in lung adenocarcinoma cells,” American Journal of Respiratory Cell and Molecular Biology, vol. 42, no. 3, pp. 320–330, 2010.

[64] J. B. Rubin et al., “Sex differences in cancer mechanisms,” Biology of Sex Differences, vol. 11, pp. 1–29, 2020.

[65] R. Ruch, “Gap Junctions and Connexins in Cancer Formation, Progression, and Therapy,” (in eng), Cancers, vol. 12, no. 11, p. 3307, 2020, doi: 10.3390/cancers12113307.

[66] S. Ylermi, “Tight junctions in lung cancer and lung metastasis: a review,” International journal of clinical & experimental pathology, vol. 5, no. 2, p. 126, 2012.

[67] A. A. Bhat et al., “Tight junction proteins and signaling pathways in cancer and inflammation: a functional crosstalk,” Frontiers in physiology, vol. 9, p. 1942, 2019.

[68] A. M. Gonzalez Guerrico, Z. M. Jaffer, R. E. Page, K.-H. Braunewell, J. Chernoff, and A. J. P. Klein-Szanto, “Visinin-like protein-1 is a potent inhibitor of cell adhesion and migration in squamous carcinoma cells,” (in eng), Oncogene, vol. 24, no. 14, pp. 2307–2316, 2005/03/31/ 2005, doi: 10.1038/sj.onc.1208476.

[69] Y. K. Chae et al., “Overexpression of adhesion molecules and barrier molecules is associated with differential infiltration of immune cells in non-small cell lung cancer,” Scientific Reports, vol. 8, 2018/01/18/ 2018, doi: 10.1038/s41598-018-19454-3.

[70] H. Y. Liu et al., “lncRNA SLC16A1-AS1 as a novel prognostic biomarker in non-small cell lung cancer,” (in eng), J Investig Med, vol. 68, no. 1, pp. 52–59, 01 2020, doi: 10.1136/jim-2019-001080.

